# Universal Exome Sequencing in Critically Ill Adults: A Diagnostic Yield of 25% and Race-Based Disparities in Access to Genetic Testing

**DOI:** 10.1101/2024.03.11.24304088

**Authors:** Jessica Gold, Colleen M. Kripke, Regeneron Genetics Center, Penn Medicine BioBank, Theodore G. Drivas

## Abstract

Numerous studies have underscored the diagnostic and therapeutic potential of exome or genome sequencing in critically ill pediatric populations. However, an equivalent investigation in critically ill adults remains conspicuously absent. We retrospectively analyzed whole exome sequencing (WES) data available through the PennMedicine Biobank (PMBB) from all 365 young adult patients, aged 18-40 years, with intensive care unit (ICU) admissions at the University of Pennsylvania Health System who met inclusion criteria for our study. For each participant, two Medical Genetics and Internal Medicine-trained clinicians reviewed WES reports and patient charts for variant classification, result interpretation, and identification of genetic diagnoses related to their critical illness.

Of the 365 individuals in our study, 90 (24.7%) were found to have clearly diagnostic results on WES; an additional 40 (11.0%) had a suspicious variant of uncertain significance (VUS) identified; and an additional 16 (4.4%) had a medically actionable incidental finding. The diagnostic rate of exome sequencing did not decrease with increasing patient age. Affected genes were primarily involved in cardiac function (18.8%), vascular health (16.7%), cancer (16.7%), and pulmonary disease (11.5%). Only half of all diagnostic findings were known and documented in the patient chart at the time of ICU admission. Significant disparities emerged in subgroup analysis by EHR-reported race, with genetic diagnoses known/documented for 63.5% of White patients at the time of ICU admission but only for 28.6% of Black or Hispanic patients. There was a trend towards patients with undocumented genetic diagnoses having a 66% increased mortality rate, making these race-based disparities in genetic diagnosis even more concerning. Altogether, universal exome sequencing in ICU-admitted adult patients was found to yield a new definitive diagnosis in 11.2% of patients. Of these diagnoses, 76.6% conferred specific care-altering medical management recommendations.

Our study suggests that the diagnostic utility of exome sequencing in critically ill young adults is similar to that observed in neonatal and pediatric populations and is age-independent. The high diagnostic rate and striking race-based disparities we find in genetic diagnoses argue for broad and universal approaches to genetic testing for critically ill adults. The widespread implementation of comprehensive genetic sequencing in the adult population promises to enhance medical care for all individuals and holds the potential to rectify disparities in genetic testing referrals, ultimately promoting more equitable healthcare delivery.

## Introduction

Technological innovations have increased the accessibility and clinical utility of broad, non-targeted genetic testing.^1^ No longer constrained by prohibitive cost or lengthy turn-around times, genomic testing, including whole exome sequencing (WES) or whole genome sequencing (WGS), is quickly becoming an integral test in the care of critically ill pediatric patients. Numerous studies of neonatal and pediatric intensive care units, including randomized control trials, show that rapid (turnaround time 1-2 weeks) or ultrarapid (turnaround time 2 days) WES/WGS increase diagnostic yield, shorten the diagnostic odyssey, and demonstrate wide-ranging healthcare cost savings due to quicker initiation of targeted treatments, shorter hospital admissions, and fewer invasive interventions.^2–8^ Expansion of rapid genomic testing to all critically ill pediatric patients replicates these clinical and financial implications.^9–11^ However, despite the well-documented benefits in pediatric populations, genomic testing is not routinely offered during the care of critically ill adults.

In the few retrospective studies of genomic testing in intensive care units (ICUs) that have included small numbers of adult patients (n=7, 36), diagnostic yields in patients over the age of 18 years ranged from 22-57% and there was no statistical difference in diagnosis based on patient age.^12,13^ The omission of adults from broad sequencing studies of critically ill patients parallels the lack of evidence-based guidance on the indications for broad genomic testing in adults more generally, despite numerous studies showing similar results to pediatric populations.^14–19^ In these studies, the diagnostic rate of exome sequencing in adults with suspected genetic conditions has been reported to range from 14-29%, demonstrating the utility of broad genetic testing approaches in this population. Together these facts suggest that incorporating rapid genomic sequencing into the care of critically ill adults may yield diagnostic rates and benefits in cost and outcome similar to those seen in the pediatric populations.

Here, we present a retrospective cohort study of outcomes of universal exome sequencing in all 365 adult patients, aged 18-40 years, admitted to any ICU of the tertiary care University of Pennsylvania Health System (UPHS) with genomic data available through the PennMedicine BioBank (PMBB). We integrated electronic health record (EHR) information, chart review, and genetic variant information to assess results by self-identified race/ethnicity and by patient knowledge of their genetic diagnosis. We identified diagnostic results in nearly 25% of critically ill adults and concerning variants of uncertain significance (VUSs) in a further 11% of patients, with most results occurring in medically actionable genes. The incidence of diagnostic variants was equivalent across all races/ethnicities and did not decrease with increasing patient age. However, Black and Hispanic patients were significantly less likely to have their genetic diagnosis documented in their medical charts, a concerning disparity given a 66% increase in mortality observed for patients with undocumented genetic diagnoses. Overall, our study suggests that the benefits of genomic testing in critical illness is age-independent, and that the universal use of broad genetic testing modalities in the critically ill adult population might improve healthcare outcomes for all patients, regardless of age, race, or ethnicity.

## Results

### Cohort Demographics and Characteristics

Of the 43,612 PMBB participants with exome sequencing data, 365 met inclusion criteria for our study (**Figure 1A**, Methods). Altogether, 48 patients (13.2%) were age 18-25 years, 92 (25.2%) were age 25-29 years, 116 (31.8%) were age 30-34 years, and 109 (29.9%) were age 35-40 years at the time of first ICU admission (**Figure 2A, Table S1**). Patients were roughly evenly divided by sex, with 200 patients (54.8%) identifying as female and 165 (45.2%) identifying as male (**Figure 2B**). Examining the EHR-recorded race and ethnicity of the 365 patients in our study, 222 (60.8%) identified as White non-Hispanic, 102 (27.9%) identified as Black non-Hispanic, 22 (6%) identified as Hispanic/Latino, 8 (2.2%) identified as Asian, and 11 (3%) identified as an other race/ethnicity (**Figure 2C**). The overall mortality rate for the study cohort was relatively low, with 30 patients (8.2%) dying during or after their hospital admission (**Figure 2D**). Comparing the broad indications for ICU admission for the study cohort (**Figure 2E**), we found that the most common diagnostic categories for ICU admission were: cardiac indications (n=77, 21.1%), cancer-related indications (n=71, 19.5%), vascular indications (n=46, 12.6%), infectious indications (n=25, 6.8%), immunologic indications (n=20, 5.5%), and renal indications (n=19, 5.2%).

**Figure 1.**
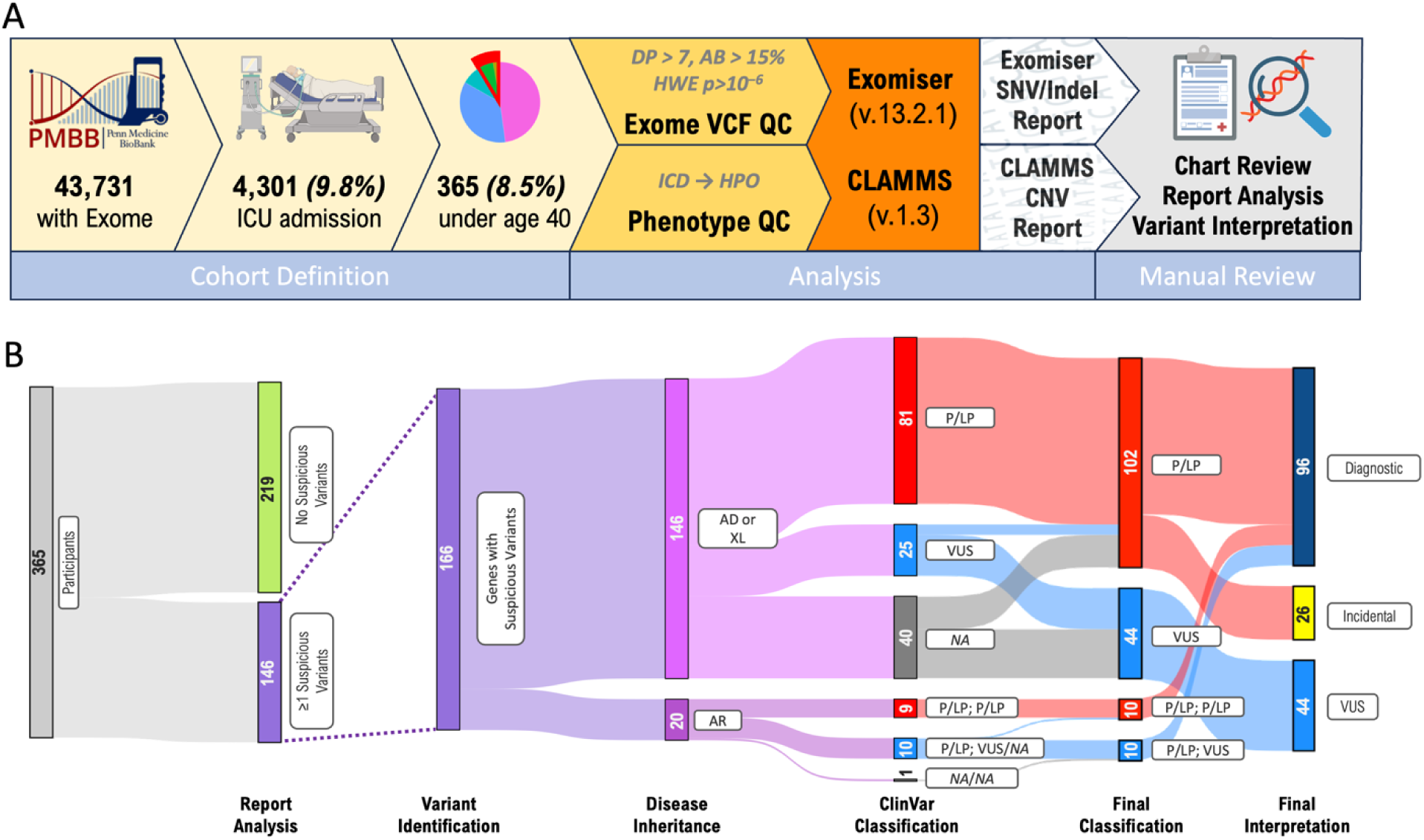
Cohort Definition and Variant Interpretation. In panel A, the overall workflow for cohort definition, report generation, and chart review is shown. Starting with the 43,731 PMBB participants with exome sequencing data, we took the subset of individuals under the age of 40 who had ever been admitted to any University of Pennsylvania Health System (UPHS) ICU with any International Statistical Classification of Diseases (ICD)-9/10 admission diagnosis code other than those falling under the category of “Injury, poisoning and certain other consequences of external causes” or “External causes of morbidity and mortality,” leaving 365 individuals in our cohort. Exome Variant Call Format (VCF) files for each participant were subjected to quality control (QC) as shown, and all ICD codes ever associated with each participant’s medical record were extracted and mapped to human phenotype ontology (HPO) terms as described in the methods section. These data were supplied as inputs to Exomiser v.12.2.1 and CLAMMS v.1.3 to generate an Exomiser and copy number variant (CNV) report, which were subsequently reviewed by two Clinical Geneticists, both board certified in Internal Medicine and Medical Genetics, with concomitant review of the patient chart. In panel B, the workflow and results of variant identification and interpretation is shown. Of the 365 participants, 146 had one more more suspicious variants identified on either/both the Exomiser or CNV report. We classified these variants by their associated inheritance pattern and by ClinVar classification, if available. For all ClinVar-annotated VUSs, and for variants absent form ClinVar, we classified variants using the American College of Medical Genetics and Genomics (ACMG) guidelines for clinical sequence interpretation. The resulting variants were subsequently classified as either diagnostic, incidental, or VUS, as described in the methods section.

**Figure 2.**
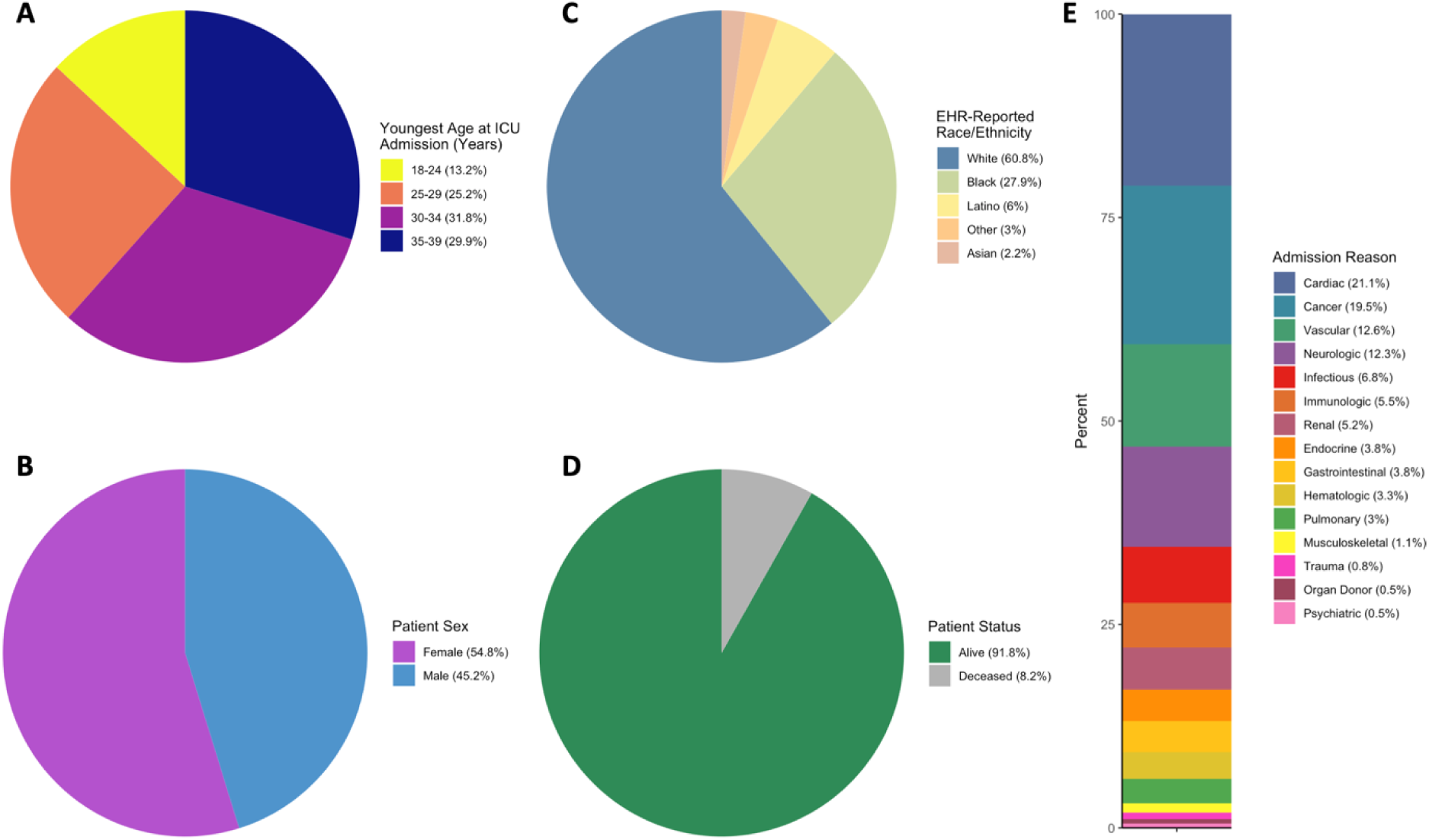
Demographics of the Study Cohort. Overall demographics of the 365 participants included in our study are shown. Panel A shows percent of the study population falling into each of four age groups at the time of earliest ICU admission. Panel B shows the percent of participants by EHR-reported patient sex. Panel C shows the percent of participants by EHR-reported race and ethnicity. Panel D shows the percent of participants by patient status (deceased or alive) at the time of discharge. Panel E shows the percent of participants divided by reason for ICU admission, broadly divided into 15 indication groups.

Broadly dividing participants into three groups by race/ethnicity – White (n=222), Black/Hispanic (individuals identifying as Black and/or Hispanic/Latino, n=124), and Other (individuals identifying as any other race/ethnicity, n=19), we found no significant demographic differences (**Table S2**). Specifically comparing the two largest groups, White and Black/Hispanic, we found no significant differences in age, sex, or patient status (deceased vs. alive). We did observe two nominally significant differences in ICU admission indications, with Black/Hispanic patients being significantly more likely to be admitted for infectious symptoms (p=0.0172) and less likely to be admitted for cancer-related issues (p=0.009). There were no significant differences in any of the other 13 ICU admission indications examined.

### Variant Identification and Interpretation

After review of medical records and the CNV and Exomiser^20^ reports generated from exome sequencing data, 187 suspicious genetic variants affecting 166 genes were identified across 146 of the 365 individuals (40.0%, **Figure 1B, Table S1**). Of these variants, 77 (41.2%) were missense variants, 36 (19.3%) were frameshift variants, 24 (12.8%) were nonsense variants, 23 (12.3%) were variants predicted to affect mRNA splicing, 18 (9.6%) were in-frame deletions or insertions, six (3.21%) were large deletions spanning more than one gene, and one each (0.5%) were start loss, stop loss, or synonymous variants. Of the 187 suspicious variants identified, 143 (76.5%) across 107 individuals (29.3% of all 365 individuals studied) were found to affect genes causative of autosomal dominant disease, three variants (1.6%) across three individuals (0.8%) were found to affect genes causative of X-linked disease, and 41 variants (21.9%) across 20 individuals (5.5%) were found to affect genes causative of autosomal recessive disease. No suspicious variants affecting the mitochondrial genome were identified.

Assessing variant pathogenicity (**Figure 1B, Table S1**), we found that 111 of the 187 suspicious variants identified (59.35%) had previously been annotated in ClinVar^21^ as pathogenic or likely pathogenic, 31 (16.58%) had previously been annotated as variants of uncertain significance (VUSs), and 47 (25.13%) had not been previously annotated in ClinVar at all. Following the American College of Medical Genetics and Genomics (ACMG) guidelines for clinical sequence interpretation,^22^ we reassessed all ClinVar-annotated VUSs: we reclassified six of the 31 VUSs (19.4%) as likely pathogenic while the remaining 25 remained classified as VUSs. Of the 47 variants lacking ClinVar annotations, we classified 20 (42.6%) as pathogenic/likely pathogenic while the remaining 27 (57.5) were classified as VUSs. None met criteria to be classified as benign/likely benign.

Exome sequencing results identifying pathogenic/likely pathogenic variants were considered “diagnostic” if the result was considered relevant to the individual’s ICU admission; otherwise, pathogenic results were classified as “incidental.” All VUS results were classified as “VUS.” Altogether, we identified 95 diagnostic results, 27 incidental findings, and 44 VUS results across the 365 participants in the study (**Figure 1B, Table S1**). While 127 individuals (34.8% of the 365 studied) had a single finding identified (either diagnostic, incidental, or VUS), 19 individuals (5.2%) had more than one finding: four individuals (1.1%) were found to have two diagnostic findings related to their admission; eight individuals (2.2%) were found to have one diagnostic finding and one or more incidental findings; three individuals (0.8%) were found to have one diagnostic result and one suspicious VUS; two individuals (0.6%) were found to have two suspicious VUSs; and one individual each (0.3%) was found to have one suspicious VUS and one incidental finding, and two incidental findings.

### High Diagnostic Rate of Universal Exome Sequencing in Adult ICU Patients

Across all 365 individuals included in our study, 90 (24.7%) were found to have one or more diagnostic results relevant to their ICU admission (**Figure 3A**). An additional 40 (11.0%) were found to have one or more suspicious VUSs, and an additional 16 (4.4%) were found to have one or more incidental findings. Overall only 219 individuals (60%) had completely negative exome results. We observed no significant differences in diagnostic or VUS rates across different racial/ethnic groups, although White individuals had the highest observed diagnostic rate and lowest observed VUS rate. The diagnostic rate in White individuals was found to be 27.0%, 21.0% in Black/Hispanic individuals, and 21.1% in individuals of any other race/ethnicity (**Figure 3B**). The rate of VUS identification in these same groups was 10.4%, 12.1%, and 10.5% respectively.

**Figure 3.**
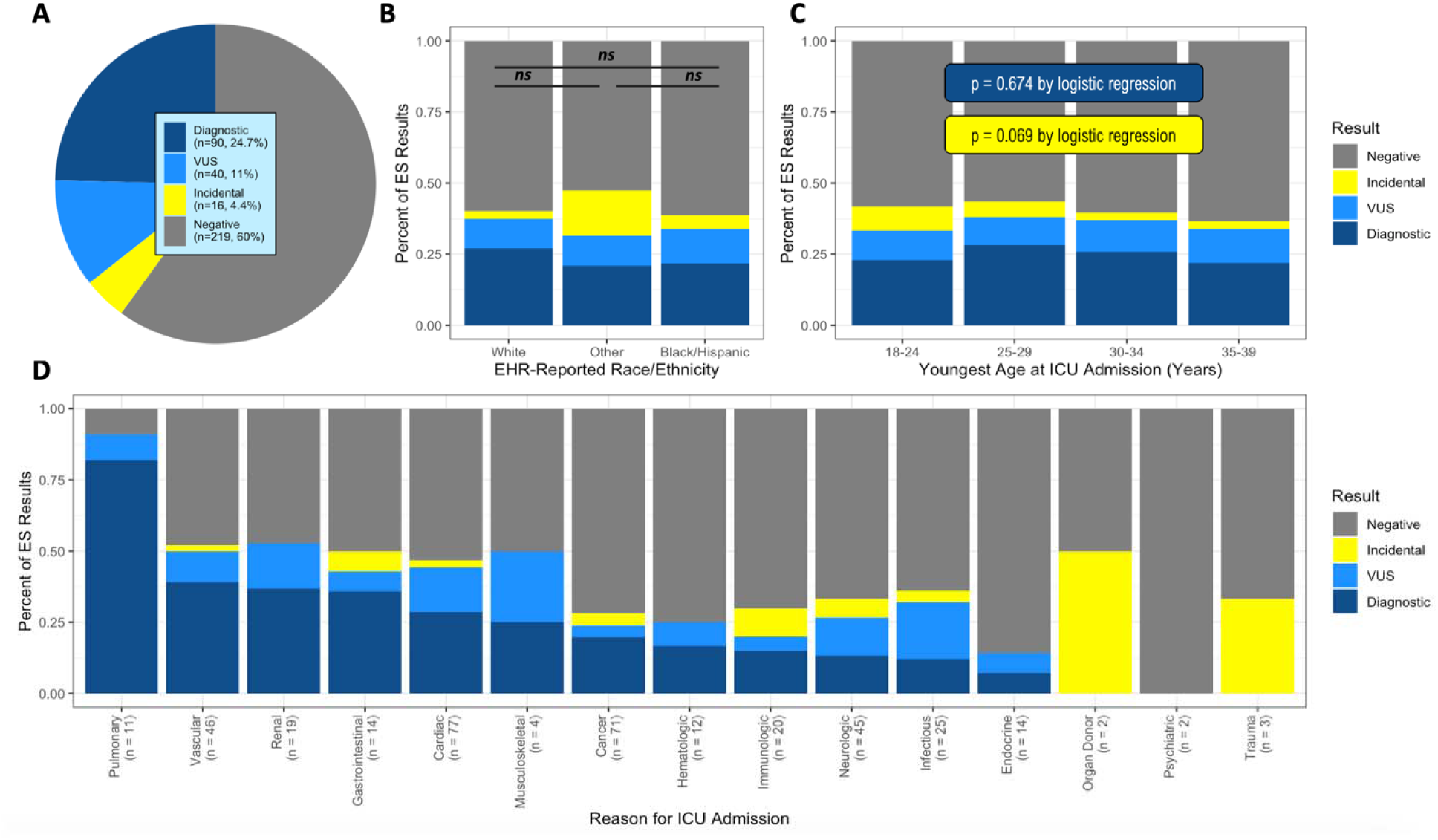
Results of universal exome sequencing in critically ill adults age 18-40 years. (A) Overall diagnostic rate of exome sequencing in the complete cohort of 365 participants, along with the rate of VUS identification and incidental diagnosis identification. In this figure each participant is counted only once, only for the highest order finding discovered on exome sequencing, with diagnostic results being evaluated first, VUSs second, and incidental finding last. (B) Overall diagnostic rate of exome sequencing as in panel A, but stratified by EHR-reported race/ethnicity. No significant differences in the rate of diagnostic findings was observed by logistic regression between any of the three groups. (C) Overall diagnostic rate of exome sequencing as in panel A, but stratified by patient age at youngest ICU admission. No significant correlation was found between patient age and diagnostic rate or VUS rate by logistic regression. (D) Overall diagnostic rate of exome sequencing as in panel A, but stratified by the reason for ICU admission, as in Figure 2E.

### Diagnostic Rate of Exome Sequencing Does Not Decrease with Increasing Patient Age

Strikingly, and contrary to other published studies,^13–15,23^ we did not observe any significant decrease in the diagnostic rate of exome sequencing in adult patients with increasing patient age (p=0.674 by logistic regression, **Figure 3C**). The diagnostic rate in patients aged 18-24 years was 22.9%, 28.3% for patients aged 25-29 years, 25.9% for patients aged 30-34 years, and 21.1% for patients aged 35-40 years. There was a trend towards decreasing prevalence of incidental findings with increasing patient age, with incidental findings identified in 8.33% of patients aged 18-24 years, 5.43% of patients aged 25-29 years, 2.59% of patients aged 30-34 years, and 3.67% in patients aged 35-40 years, although this did not reach statistical significance (p = 0.069).

### Highest Yield of Exome Sequencing in Pulmonary, Vascular, and Renal Disease Patients

Dividing patients by indication for ICU admission, we found that certain indications were more likely to yield diagnostic results than others (**Figure 3D**). The five admission indication groups with the highest diagnostic rate were pulmonary disease (n=11, diagnostic rate 81.8%), vascular disease (n=46, diagnostic rate 39.1%), renal disease (n=19, diagnostic rate 36.8%), gastrointestinal disease (n=14, diagnostic rate 35.7%), and cardiac disease (n=77, diagnostic rate 28.6%). The lowest diagnostic rates were observed for infectious diseases (n=25, diagnostic rate 12.0%), endocrine disease (n=14, diagnostic rate 7.1%), and organ donor status, psychiatric disease, and trauma, all of which had a diagnostic rate of 0% (n=2, 2, 3, respectively).

### Cardiac, Vascular, and Malignancy-Associated Genes Predominate

Greater than 50% of all diagnostic results identified affected genes causing cardiac (n=18, 18.9%), cancer (n=16, 16.8%), or vascular (n=16, 16.8%) phenotypes (**Figure 4A**). The same was true for VUSs (cardiac n=17, 38.6% of VUSs; cancer n=2, 4.55%; vascular n=4, 9.09%). While most genes were only represented once among the diagnostic results (n=48, 51%), 13 were diagnosed more than once (**Figure 4B**). These include *FBN1* (n=8), *CFTR* (n=7), *TTN* (n=6), *BRCA2* (n=4), *VHL* (n=4), *LMNA* (n=3), *PKD1* (n=3), and *ACVRL1*, *CACNA1S*, *MYH7*, *NF1*, *PLN*, and *SMAD3* (n=2 each). No genes appeared more than once in the VUS dataset. In the dataset of incidental findings, the majority of genes were identified only once (n=16, 59%, **Figure 4C**), while five genes appeared at least two times: *ALPL* (n=3), and *BRCA2*, *MEFV*, *PALB2*, and *TTN* (n=2 each).

**Figure 4.**
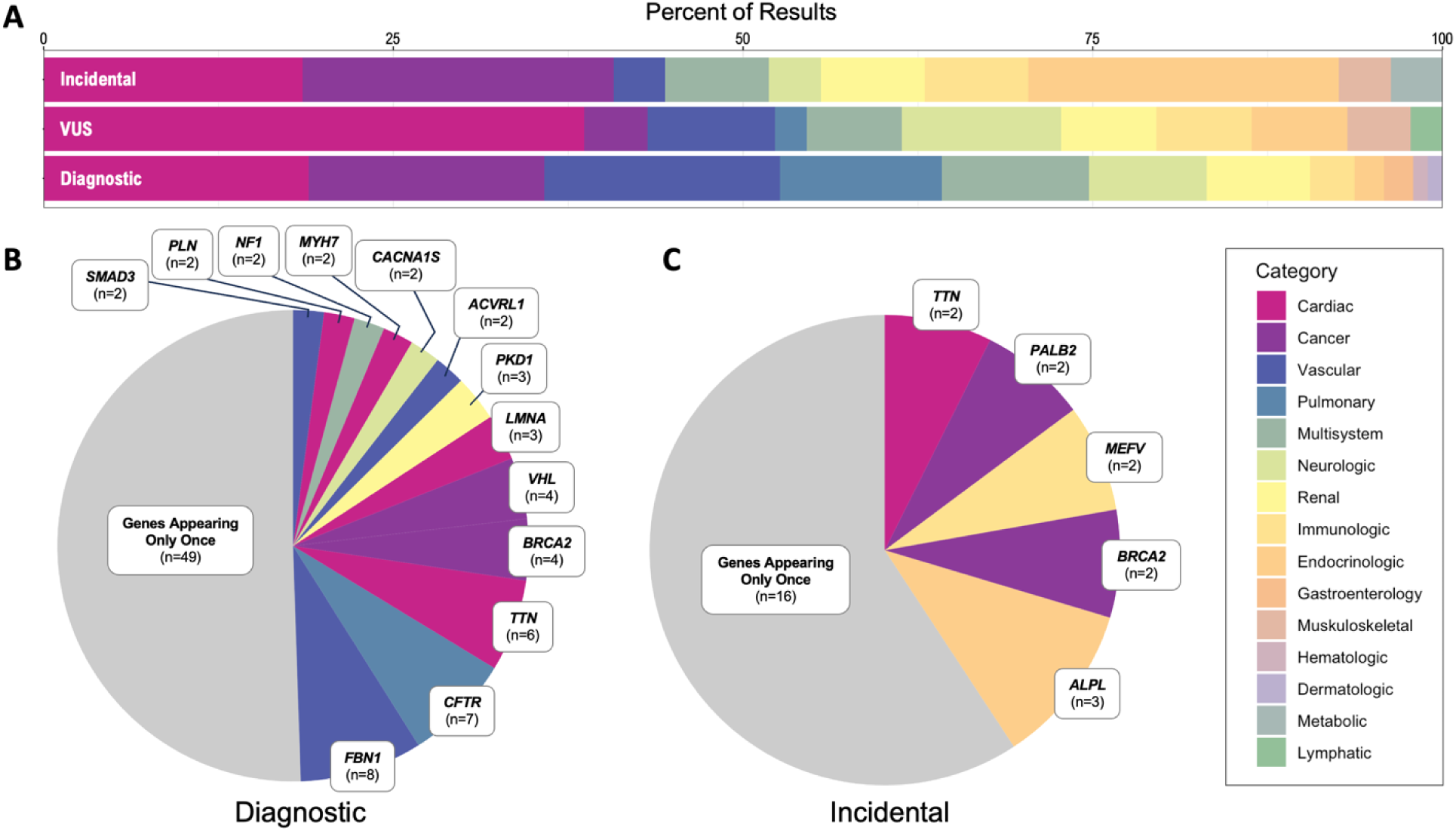
Distribution of genes identified by exome sequencing across different result categories. (A) Proportion of exome sequencing results colored by the diagnostic category of the involved gene. Results are stratified by diagnostic, VUS, and incidental results. (B) Frequency of gene implicated in diagnostic findings. Genes identified only once within this dataset are collectively labeled as “Genes Appearing Only Once.” The remaining genes, appearing two or more time across all diagnostic results, are displayed. Color coding corresponds to the categories defined in panel A. (C) Frequency of genes implicated in incidental findings. Genes identified only once within this dataset are collectively labeled as “Genes Appearing Only Once.” The remaining genes, appearing two or more times across all incidental results, are displayed. Color coding corresponds to the categories defined in panel A.

### Half of All Diagnostic Results Are Unknown/Undocumented in Patient Charts

For the 95 diagnostic exome results we identified, chart review revealed that many were unknown or undocumented in the patient chart at the time of ICU admission (**Figure 5A**). Overall, 44 of the 95 diagnoses were absent from patient charts (46.3%). There was a trend towards older patients being less likely to have a documented diagnosis; diagnoses were documented for 58.3% of patients aged 18-24 years, 59.3% of patients aged 25-29 years, 62.5% of patients aged 30-34 years, but only for 33.3% of patients aged 35-40 years, however this relationship with age was not statistically significant by logistic regression (p = 0.232, **Figure 5B**).

**Figure 5.**
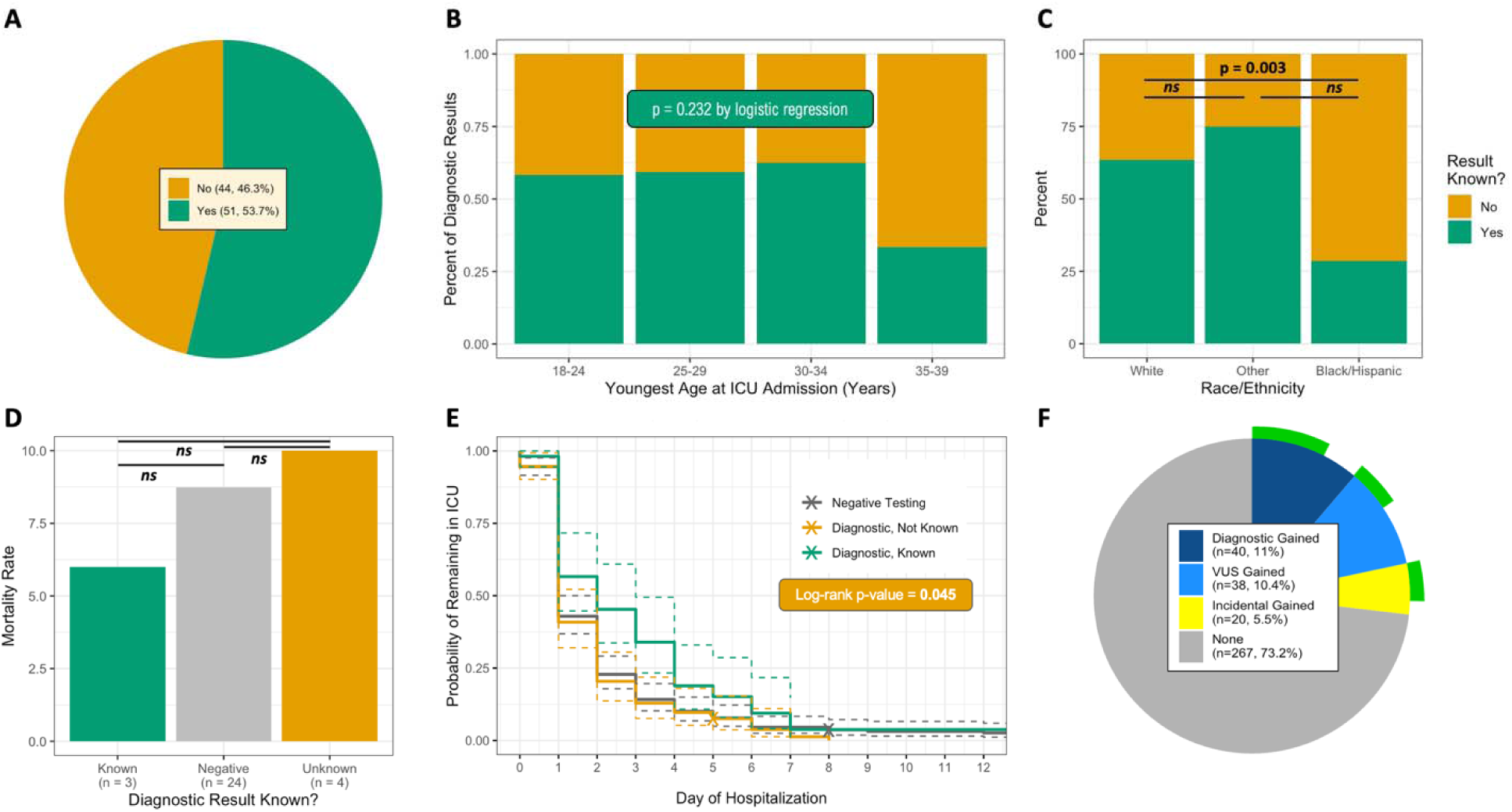
Racial/ethnic disparities in documentation of genetic diagnoses and implications for clinical outcomes. (A) Percent of all 95 diagnostic results identified on exome sequencing which were known and documented in patient charts prior to ICU admission (yes, in green) or unknown/undocumented in patient charts (no, orange). (B) Percent of diagnostic exome results known and documented, as in panel A, but stratified by patient age. No significant correlation was found between patient age and documentation of genetic diagnosis by logistic regression. (C) Percent of diagnostic exome results known and documented, as in panel A, but stratified by EHR-recorded race/ethnicity. Black patients and Hispanic patients are significantly less likely to have a documented genetic diagnosis compared to White patients by logistic regression. (D) Mortality rate for all 365 patients included in our study, stratified by result of exome sequencing (diagnostic vs. negative) and by documentation of results in the patient chart. Patients with documented diagnostic results were less likely to die compared to patients with negative exome results, who were less likely to die compared to patients with undocumented diagnostic results, although these difference were not statistically significant by logistic regression. (E) Kaplan-Meier curve illustrating the likelihood of remaining in the ICU by day of hospitalization, stratified into the same three groups as in panel D. Patients with documented diagnostic results had significantly longer lengths of stay in the ICU by log-rank test. (F) Percent of patients that would gain a new diagnosis not already documented in their electronic medical record, stratified by diagnostic category, across all 365 patients included in the study. Green bars indicate the proportion, within each diagnostic category, that would specifically gain a diagnosis in a gene with clearly defined management recommendations.

### Black/Hispanic Patients are Significantly Less Likely to Have Documented Diagnoses

Comparing the rates at which diagnostic results were known and documented by race/ethnicity, significant differences emerged (**Figure 5C**). White patients were significantly more likely to have a documented diagnosis compared to Black/Hispanic patients (p=0.003 by logistic regression). Diagnoses were documented for 63.5% of White patients and 75% of patients of other race/ethnicity, but only for 28.6% of Black/Hispanic patients. This disparity cannot be explained by demographic differences (**Table S2**), or differences in overall diagnostic rate of exome sequencing between these different groups (**Figure 3B**).

### Patients with Unknown/Undocumented Diagnoses Have Higher Mortality Rates

The overall mortality rate for participants in our study was thankfully relatively small (8.2%, **Figure 2D**), but we observed differences in mortality rate by diagnostic status (**Figure 5D**). Overall, 6.0% of patients with chart-documented diagnostic results died either during or after their hospitalization, compared to 8.7% of patients with negative exome results and 10.0% of patients with diagnostic but undocumented exome sequencing results. These differences did not reach statistical significance, but the 66% increase in mortality rate observed for patients with undocumented diagnostic results compared to patients with documented diagnostic results is notable.

### Patients with Documented Diagnoses Have Significantly Longer ICU Admissions

We next examined the length of stay (LOS) in the ICU for our cohort, divided by diagnostic status. Patients with chart-documented diagnostic results had a mean ICU LOS of 3.0 days (median 2 days), compared to 2.2 days (median 1 day) for patients with negative exome results and 1.9 days (median 1 day) for patients with diagnostic but undocumented diagnostic exome sequencing results. The difference in LOS observed for patients with undocumented diagnostic results compared to patients with documented diagnostic results was significant by ANOVA (p=0.027). Generating Kaplan-Meier curves to compare the LOS between groups by survival analysis the same significant different was seen by log-rank test (p=0.045, **Figure 5E**).

### Universal exome sequencing reveals medically actionable results in 8% of patients

Excluding patients with known diagnoses, 41 of the 365 individuals that we studied (11.2%) would have gained a diagnostic result from universal exome sequencing, 38 (10.4%) would have gained a diagnosis of a suspicious VUS, and 19 (5.2%) would have gained a diagnosis of an incidental pathogenic finding (**Figure 5F**). Of all of the 166 genetic findings we identified in our study, 108 (65.1%) occurred in genes with specific management guidelines described in the NCBI resource “GeneReviews” (**Table S1**).^24^ For diagnostic results, 72 (76.6%) occurred in genes with medically management recommendations; for incidental findings 17 occurred in medically actionable genes (60.7%), and for VUS results 19 (43.2%) occurred in genes with medically management recommendations. For diagnostic results that were specifically known/documented in the patient chart 42 (82.4%) occurred in genes with clearly defined medical management guidelines, while for results that were not known/documented 30 (69.8%) occurred in medically actionable genes. This suggests that for 30 patients (8.2% of our entire cohort), exome sequencing at the time of admission might have suggested specific management changes.

## Discussion

Our study is, to our knowledge, the first of its kind to investigate the utility of broad exome sequencing in the critically ill young adult population. The results serve as a pivotal advancement in our understanding of the impact of Mendelian disease in adults, highlighting the frequency of genetic contribution to critical illness in this understudied population. We find that nearly a quarter of all adult UPHS ICU patients aged 18-40 have a Mendelian genetic diagnosis related to their critical illness. For over 75% of patients, these genetic diagnoses confer specific care-altering management recommendations. Notably, we find consistently high yield of exome sequencing across all age brackets examined, even in this patient population unselected for enrichment of suspected genetic diagnoses. These results highlight the fact that genetic testing is not merely relevant to the young or to those with *a priori* suspicions of a genetic disorder. The implication here is profound: there exists an untapped potential for genetic testing that could benefit a much broader adult demographic than previously recognized, altering medical management for a large number of critically ill patients. These findings stand to reshape our approach to genetic assessments in the adult patient population, which has all too often been overlooked for the implementation of broad genetic testing.

Our findings stand in contrast to the traditionally held view that the likelihood of uncovering genetic diagnoses decreases with increasing patient age but is in keeping with other studies of the yield of exome/genome sequencing in adults. In a study of whole genome sequencing in 100 generally healthy patients aged 40–65 years in the primary care setting, Vassy *et al*. discovered a new genetic diagnosis in 22%,^16^ although many patients exhibited only minimal symptoms of their diagnosis. In other studies of adult patients with suspicion for a genetic diagnosis, ^14,15,17–19^ the diagnostic rate of exome sequencing has been reported to range from 14-29% – lower than that observed in the pediatric population,^25^ but high enough to demonstrate the utility of broad genetic testing approaches in adults. The diagnostic rate of 24% that we report in our study is in keeping with these previous reports, despite the fact that the population we examined was not specifically selected for suspicion of genetic disease. This suggests that the presence of critical illness in a young adult is as good a predictor of Mendelian disease as “suspicion for a genetic disease” as determined by a genetics-trained provider.

The high yield of broad exome sequencing that we report in critically ill adults can be put in the context of a number of recent similar studies of broad exome/genome sequencing in the critically ill pediatric and neonatal patient populations that have reported diagnostic rates of 21-38%.^2–4,9,10^ The diagnostic yield that we report here is essentially the same as those reported in these studies, suggesting that critical illness is a strong predictor of Mendelian disease across the life span. Importantly, many of these studies found significant cost savings and important changes in management due to rapid exome/genome sequencing. This, along with our results identifying specific medical management recommendations for over 75% of identified genetic diagnoses, suggests that rapid exome/genome sequencing in critically ill adults might prove similarly beneficial at improving patient care and lowering healthcare costs, although prospective studies are needed to validate this hypothesis.

In nearly half of all patients with diagnostic exome sequencing results, we find that this diagnosis is unknown to the patient or their treatment team. We also found that patients with documented genetic diagnoses had significantly longer ICU stays and trended towards having lower mortality rates than patients with unknown/undocumented genetic diagnoses. We cannot be certain why the presence of a known genetic diagnosis was associated with significantly longer ICU length of stay, but it is possible that these patients are treated more cautiously than their counterparts without known genetic disorders, and it is possible that this might explain the lower mortality rate that we observed in this group. Future prospective studies are needed to understand how critical care management changes in the face of a genetic diagnosis, and whether making such a diagnosis truly decreases patient mortality. This consideration is not trivial; the shift toward precision medicine could herald a more efficient allocation of resources and a reduction in healthcare expenditures by tailoring interventions to individual genetic profiles, leading to improved outcomes and more cost-effective care.

One of the most disconcerting revelations of our study is the stark disparity in the awareness of genetic diagnoses along racial and ethnic lines. We found no difference in the diagnostic rate of exome sequencing between White and Black/Hispanic patients; however, our data unambiguously show that Black and Hispanic patients are substantially less likely to have their genetic diagnosis known and documented compared to their White counterparts. This is alarming in the context of the 66% increased mortality rate we observed in patients with undocumented genetic diagnoses. This is more than a statistic; it is indicative of a systemic failure in the delivery and implementation of clinical genetic testing and precision medicine that potentially contributes to the significant healthcare disparities observed in non-White patients.^26,27^ The reality that a patient’s race or ethnicity could influence their likelihood of receiving a timely and accurate genetic diagnosis is a glaring call to action. It demands an interrogation into the access, education, and biases that pervade our healthcare systems, and suggests that broad, universal testing approaches such as those examined in our study may be needed to eradicate such inequities.

One striking feature of our results was the high rate at which we repeatedly identified pathogenic variants in the same gene in different patients, with half of all diagnostic results affecting genes appearing more than once in our data. Five genes alone – *FBN1*, *CFTR*, *TTN*, *BRCA2*, and *VHL* – were found to underly the diagnoses of 29 patients included in our study, representing 30% of all diagnostic results. This is in contrast to what has been observed in studies of broad sequencing in pediatric ICU populations, where recurrent diagnoses make up only a small portion of the diagnostic results.^2,3^ Additionally, the recurrent genes we identify are not overlapping with the few recurrent genes identified in these pediatric populations. This suggests that the genetic landscape of Mendelian disease in critically ill adults may be qualitatively different, and potentially more stereotyped, than what is observed in pediatric population. However, it is also possible that the specific expertise offered at the tertiary care hospital system that we studied directly influenced this finding, with certain diseases and phenotypes being specifically enriched in the patient population of UPHS. For example, the large and well regarded Aorta Center at UPHS attracts patients from around the country with aortic aneurysmal disease^28^ and may have resulted in the large number of patients with *FBN1* pathogenic variants identified in our study.

Furthermore, and similar to the above point, the vast majority of diagnoses we identified result in diseases of adult onset. We did not exclude patients with known genetic diagnoses from our study, and thus the conspicuous scarcity of pediatric-onset genetic syndromes in our adult ICU cohort raises pressing questions about the intersection of genetics with patient survival and healthcare delivery. Are individuals with pediatric-onset genetic conditions less likely to survive into adulthood? Are they not experiencing critical illness in adulthood? Are they still seeking critical care treatment at pediatric hospitals? Is clinician bias discouraging ICU-level treatment?^29^ Are they not enrolling in the PMBB, a research-based cohort that requires specific consent, and thus not captured by this study? The answers to these questions remain elusive and clearly more research is needed to understand this discrepancy. However, it is clear that a re-evaluation of our approach to genetic testing throughout a patient’s lifespan is needed, with the need for a dynamic model that adapts to the evolving clinical presentation of genetic disease across the lifespan. Genetic diseases present differently in adulthood, often with symptoms that are not present in childhood and adolescence, and the same care models and approaches that have been proven effective in pediatric populations may not be appropriate for adult patients with genetic disease.

In considering the broader implications of our study, it is paramount to acknowledge its limitations. First and foremost, this was a single-center study, which comes with a number of limitations and potential confounders as discussed above. Secondly, this was a retrospective study that utilized research-grade sequencing data. The retrospective nature of the study makes it impossible for us to know how a genetic diagnosis might have affected patient care in real-time, and it is possible that some of our sequencing results, completed by a non-CLIA-certified sequencing lab, may be spurious. That being said, previous studies have shown CLIA validation of research-grade biobank exome sequencing to exceed 99%.^30^ Lastly, a major limitation of our work is that it depends entirely on patient enrolment in PMBB. Although overall demographics are not significantly different between PMBB and UPHS,^31^ it is possible that the population we studied represents a subset of patients that is principally different from the overall UPHS ICU-admitted patient population. These limitations necessitate cautious interpretation of the findings and underscore the need for prospective, multicenter studies of clinical-grade exome sequencing to validate and extend our results.

In conclusion, our findings paint a picture of a medical landscape on the cusp of transformation. The evidence for the utility of universal exome sequencing in critically ill adults is compelling, with new diagnoses and potential management changes identified in more than 10% of all adult patients aged 18-40 years admitted to any ICU. As we stand on the brink of this new frontier, it is incumbent upon us to forge a path forward that not only recognizes the importance of Mendelian genetic disease in adult morbidity and mortality, but actively incorporates this knowledge into routine clinical practice to avoid exacerbating healthcare disparities and to improve healthcare outcomes across the board. It is through such endeavors that we may strive toward a future where precision medicine is not a lofty ideal but a standard component of adult critical care and a tangible and potentially life-saving reality for all patients.

## Methods

### PMBB patient recruitment and exome sequencing

The Penn Medicine BioBank (PMBB)^31^ is a University of Pennsylvania academic biobank which recruits patient-participants from the University of Pennsylvania Health System (UPHS) around the greater Philadelphia area in the United States. Appropriate consent was obtained from each participant regarding storage of biological specimens, genetic sequencing, and access to all available EHR data, and permission to recontact for future studies. The study was approved by the Institutional Review Board of the University of Pennsylvania and complied with the principles set out in the Declaration of Helsinki (protocol # 854452). This study included the subset of 43,731 individuals enrolled in PMBB who had previously undergone exome sequencing. Briefly, for each individual, DNA was extracted from stored buffy coats and exome sequences were generated by the Regeneron Genetics Center (Tarrytown, NY) and mapped to GRCh38 as previously described.^32^ For quality control (QC), sample-level filtering was as follows: individuals with low exome sequencing coverage (less than 75% of targeted bases achieving 20× coverage) or with high missingness (greater than 5% of targeted bases) were removed from analysis, leaving 43,612 samples after sample-level filtering. Variant-level filtering was as follows: in each sample, all single nucleotide variants (SNVs) with a total read depth < 7 were changed to “no-call”, and similarly all insertion/deletion (INDEL) variants with a total read depth < 10 were changed to “no-call.” Subsequently we removed any variant sites where no sample carried an alternate allele balance ≥ 15% (SNVs) or 20% (INDELs).

#### Patent cohort definition

Of the 43,612 PMBB participants with exome sequencing data, we excluded individuals with no record of admission to a UPHS intensive care unit, leaving 4,590 individuals. We next removed any individuals with ICU admission diagnosis codes belonging to ICD-10 chapters XIX (Injury, poisoning and certain other consequences of external causes) or XX (External causes of morbidity and mortality), resulting in the exclusion of 289 additional individuals. Lastly, we excluded all patient whose age at earliest recorded ICU admission was less than 18 years or greater than or equal to 40 years, leaving a total of 365 individuals for inclusion in the study.

#### Copy Number Variant (CNV) Report Generation

Copy number variants (CNVs) were annotated, starting with the same exome sequencing data as described above, using version 1.3 of the CLAMMS pipeline.^33^ Standard quality control measures were taken both at the sample level (samples with >40 CNVs or with >40,000 exons called as CNVs were removed) and chromosome level (for samples with >10% of a chromosome covered by >1 CNV call, that chromosome was removed). QC levels ranging from 0-3 were assigned to each CNV call based on Q_non_dip, Q_exact, and allele balance and heterozygosity metrics. Only CNVs meeting the most stringent QC threshold of 3 were included in the analysis. For each individual in the study, a table of the resultant CNV calls, along with affected protein coding genes, was generated for manual review as described below.

### Patient phenotype definition

For the 365 patients included in our study, all International Statistical Classification of Diseases (ICD)-9 and ICD-10 disease diagnosis codes and procedural billing codes ever associated with their care were extracted from the electronic health record (EHR). ICD-9 codes were mapped to ICD-10 using the Center for Medicare and Medicaid Services 2017 General Equivalency Mappings (https://www.cms.gov/Medicare/Coding/ICD10/2017-ICD-10-CM-and-GEMs.html) with unmappable ICD-9 codes dropped from further analysis. For each individual, each unique ICD-10 code was subsequently mapped to a Human Phenotype Ontology (HPO) code using equivalency mappings as published by McArthur et al.^34^ and the unique set of HPO terms, per individual, was passed to Exomiser as input.

#### Exomiser Report Generation

For each individual included in the study, a list of HPO terms and the QCed VCF file of exome sequencing results, both generated as described above, was passed to the tool Exomiser v.13.2.1,^20^ a bioinformatic tool for prioritizing potential disease-associated genetic variants for manual review using random-walk analysis of protein interaction networks, clinical phenotype comparison with known patients based on HPO terms, cross-species phenotype comparisons, as well as a wide range of other computational filters for variant frequency and predicted pathogenicity. Exomiser was run using default/recommended settings considering all possible inheritance patterns, incorporating variant allele frequencies from gnomAD, exAC, and TOPMED, and excluding variants annotated as affecting non-coding regions of the genome. For each individual included in our study Exomiser output in HTML formal was generated for manual review as described below.

#### Chart Review, Variant Pathogenicity Assertions, and Diagnoses

For each of the 365 individuals included in our study, two physicians, both trained and board certified in Internal Medicine and Medical Genetics, reviewed the CNV report, Exomiser report, and electronic health record of each patient to identify genetic variants that might be relevant to each patient’s ICU admission. A short clinical summary of each patient’s medical history was prepared and documented. Suspicious genetic variants deemed at last possibly related to an individual’s ICU admission by both physicians (considering both the patient phenotype, variant classification, and the disease inheritance pattern) were selected for further review. Altogether, 187 suspicious variants affecting 166 genes were identified across 146 individuals (**Figure 1B**). Using the American College of Medical Genetics and Genomics (ACMG) guidelines for clinical sequence interpretation,^22^ we classified/reclassified all variants either absent from the ClinVar database, or with ClinVar entries of VUS only. Subsequently, for all pathogenic/likely pathogenic variants, exome sequencing results were classified as being “Diagnostic” if the pathogenic variant was deemed relevant to the patient’s ICU admission and fit the appropriate inheritance pattern for the disease of interest, or classified as being “Incidental” if the identified pathogenic variant was deemed medically relevant, but not directly related to the patient’s ICU admission. For autosomal recessive diseases, results were considered diagnostic if at least one of the two variants identified was pathogenic/likely pathogenic and if the associated disease phenotype was felt to be a strong match for the patient’s presentation. All results with only VUSs identified were classified into a third group, “VUS.” Patient charts were specifically reviewed to determine if the identified genetic variant had previously been identified clinically and documented in the chart. Patients with non-molecularly confirmed clinical diagnoses were considered to have been previously diagnosed for this purpose. We subsequently classified all genes with identified variants based on the primary organ system affected, and by the presence or absence of specific medical management guidelines in the NCBI GeneReviews resource.^24^

#### Data Visualization and Statistical Analysis

All data analysis and visualization was completed in RStudio using R version 4.3.0 and the following packages: ggplot2 v.3.4.3, ggsankey v.0.0.9, dplyr v.1.1.3, tidyverse v.2.0.0, survival v.3.5. Statistical analyses were carried out, as indicated, by constructing linear/logistic regression models or by log-rank test as indicated in the figure legends. For all analyses, two-sided p-values less than 0.05 were considered nominally significant.

## Data Availability

All summary and aggregate data produced in the present study are available upon reasonable request to the authors. Individual-level genetic data cannot be shared due to concerns for patient privacy.

## Acknowledgments

We acknowledge the Penn Medicine BioBank (PMBB) for providing data and thank the patient-participants of Penn Medicine who consented to participate in this research program. We would also like to thank the Penn Medicine BioBank team and Regeneron Genetics Center for providing genetic variant data for analysis. We thank Daniel Rader and Michael P. Hart for their discussion and input on study conception and design. The PMBB is approved under IRB protocol# 813913 and supported by Perelman School of Medicine at University of Pennsylvania, a gift from the Smilow family, and the National Center for Advancing Translational Sciences of the National Institutes of Health under CTSA award number UL1TR001878. T.G.D. is supported in part by the NIH K08 1K08DK127247 and by the Burroughs Wellcome Fund.

## SUPPLEMENTAL INFORMATION

### List of Supplemental Tables

Table S1. Exome sequencing results and associated participant characteristics and demographics

Table S2. Demographic characteristics of the study cohort, divided by EHR-reported race/ethnicity

## PENN MEDICINE BIOBANK BANNER AUTHOR LIST AND CONTRIBUTION STATEMENTS

### PMBB Leadership Team

Daniel J. Rader, M.D., Marylyn D. Ritchie, Ph.D.

Contribution: All authors contributed to securing funding, study design and oversight. All authors reviewed the final version of the manuscript.

### Patient Recruitment and Regulatory Oversight

JoEllen Weaver, Nawar Naseer, Ph.D., M.P.H., Giorgio Sirugo, M.D., P.h.D., Afiya Poindexter, Yi-An Ko, Ph.D., Kyle P. Nerz

Contributions: JW manages patient recruitment and regulatory oversight of study. NN manages participant engagement, assists with regulatory oversight, and researcher access. GS assists with researcher access. AP, YK, KPN perform recruitment and enrollment of study participants.

### Lab Operations

JoEllen Weaver, Meghan Livingstone, Fred Vadivieso, Stephanie DerOhannessian, Teo Tran, Julia Stephanowski, Salma Santos, Ned Haubein, P.h.D., Joseph Dunn

Contribution: JW, ML, FV, SD conduct oversight of lab operations. ML, FV, AK, SD, TT, JS, SS perform sample processing. NH, JD are responsible for sample tracking and the laboratory information management system.

### Clinical Informatics

Anurag Verma, Ph.D., Colleen Morse Kripke, M.S. DPT, MSA, Marjorie Risman, M.S., Renae Judy, B.S., Colin Wollack, M.S.

Contribution: All authors contributed to the development and validation of clinical phenotypes used to identify study subjects and (when applicable) controls.

### Genome Informatics

Anurag Verma Ph.D., Shefali S. Verma, Ph.D., Scott Damrauer, M.D., Yuki Bradford, M.S., Scott Dudek, M.S., Theodore Drivas, M.D., Ph.D.,

Contribution: AV, SSV, and SD are responsible for the analysis, design, and infrastructure needed to quality control genotype and exome data. YB performs the analysis. TD and AV provides variant and gene annotations and their functional interpretation of variants.

